# Is Latin American brain anatomy “normal”? Construction and evaluation of a Chilean brain template

**DOI:** 10.64898/2026.11.15.26368168

**Authors:** Eduardo Cavieres, Julio Sotelo, Cecilia Besa, Marcelo E. Andia, Rodrigo Salas, Steren Chabert

## Abstract

Many neuroimaging processes use a brain reference anatomical template; the most widely used is the Montreal Neurological Institute (MNI) template. Yet studies have shown that different populations exhibit different global brain features, and the use of a unique Canadian template may introduce biases in analyses for other populations. The purpose of this work is to construct a brain template that accurately represents our local population in Chile. In addition, our project explicitly addresses another pressing challenge: the limited availability of high-field MR scanners in Latin America. In this context, we will build our template using a 0.55T MR scanner, thus ensuring not only population specificity but also methodological alignment with the technological realities of our healthcare systems. Healthy volunteers (n = 76, ages 18–60) underwent T1-weighted MRI. Images were preprocessed and iteratively registered via rigid and non-linear steps to generate a Chilean template (CL76). Evaluation involved dimensional comparisons with Canadian, Indian, Chinese, and Korean templates. Deformation metrics were estimated for an additional 23 volunteers. The Chilean template measures 166 × 132 × 100 mm^3^, smaller than the Canadian one (179 × 137 × 118 mm^3^), and has distinct proportions, particularly the width/height ratio of 1.31 versus 1.16. Deformation of a test population onto CL76 was significantly lower (p = 0.010) and less variable than that of MNI152. The top deformed regions onto CL76 or onto MNI152 differed, confirming population specificity on the template. To the best of our knowledge, this is the first brain template generated in South America; however, we do not claim to represent the entire continent—only a specific population from Chile. This work highlights the need for broader representation across diverse populations, particularly to counterbalance the current over-representation of individuals from high-income countries with established research and imaging infrastructures.

## Introduction

Many neuroimaging processes use a brain reference anatomical template, the most widely used being that of the Montreal Neurological Institute MNI152 [1,2]. This template consists of an average of 152 regular MRI scans, developed in Canada. Different MNI atlases are available, built with more subjects [3], or for different ages [4], but these templates are still developed in North America. Studies have shown that different populations exhibit differences in global brain features: this point has been observed in India with 157 subjects [5], or with 466 subjects between 6 and 60 years of age [6]; in China with 200 subjects [7] and with 674 subjects [8].

Anatomical features have been shown to be different, for example, in language-related areas in a Chinese template compared to a Caucasian template [7], or different brain growth charts have also been described [6,8]. Even though some studies include a representation of different ethnic origins [9], these templates represent mainly three continents: North America, Europe and Asia; the Latin American population is mostly absent [10]. This gap is particularly critical in clinical practice, where precise spatial localisation underpins diagnosis, lesion mapping, surgical planning, and disease monitoring. Non-representative templates risk introducing systematic biases, underscoring the need for locally derived brain templates that ensure anatomically accurate and population-relevant neuroimaging.

The purpose of this work is to construct a brain template that accurately represents our local population in Chile. Developing population-specific templates is crucial, as multiple studies have shown that the use of mismatched templates can compromise the accuracy of brain segmentation, registration, and functional activation detection [7,11,12]. This mismatch has also been demonstrated to reduce statistical power in neuroimaging studies [7,13] and to hinder the reliable detection of pathological processes such as Alzheimer’s-related atrophy [12]. Even essential preprocessing steps can be negatively affected, as illustrated by Gao et al. [14], who reported lower performance in face anonymisation when applying a Caucasian template to a Chinese cohort. These findings underscore the urgent need to expand and diversify population representation in neuroimaging resources, moving beyond Eurocentric standards.

In addition, our project explicitly addresses another pressing challenge: the limited availability of high-field magnetic resonance (MR) scanners in Latin America. While most research templates are derived from 3.0T data, such equipment is not the norm in clinical practice across the region, where lower-field systems are far more prevalent. Recent advances in low-field image acquisition and reconstruction have highlighted the potential of these technologies to improve accessibility, reduce costs, and broaden participation in neuroimaging research. In this context, we will build our Chilean brain template using a new 0.55T MR scanner, thus ensuring not only population specificity but also methodological alignment with the technological realities of our healthcare systems.

## Materials and methods

### Volunteers and images description

This study received approval from our local Ethics Committee (*Comité Ético Científico de Ciencias de la Salud de la Pontificia Universidad Católica de Chile*, ID 210125006). 99 healthy volunteers were recruited between 01/24/2023 and 08/01/2023, all providing written informed consent for their participation. Inclusion criteria were being between 18 and 60 years old and being healthy. Healthy status was based on the absence of a reported diagnosis and no medication use; psychiatric and neurological history, and vascular risk factors were not systematically recorded, which may have allowed subclinical conditions in the cohort. Subjects were excluded if their images showed abnormalities or anatomical variations greater than 3 mm: pathological findings like cysts, infarcts, or hemorrhages may introduce non-normative tissue contrasts and deformations that could bias template averaging toward abnormality, compromising spatial normalisation for downstream analyses. Anatomical variations greater than 3 mm (e.g., enlarged perivascular spaces) exceed typical inter-subject deformation residuals [15,16]; therefore, as a conservative methodological choice, we set a threshold of 3 mm to balance inclusivity of natural variability with template fidelity.

Experienced neuroradiologists reviewed the entire dataset. 23 subjects were excluded; a description of the findings is given in S1 Table. A total of 76 subjects were included in the template construction (49 women, 64% of the included population) with a mean age of 32 ± 9 years (ranging from 18 to 59 years). The average height was 1.59 ± 0.07 m for female subjects, and 1.74 ± 0.05 m for male subjects; the age and height distributions are shown in Fig 1. Average weight was 67.7 ± 14.4 kg for women and 77.8 ± 12.8 kg for men. The study group had an average body surface area of 1.80 ± 0.21 m^2^ (ranging from 1.34 to 2.31 m^2^).

**Fig 1.**
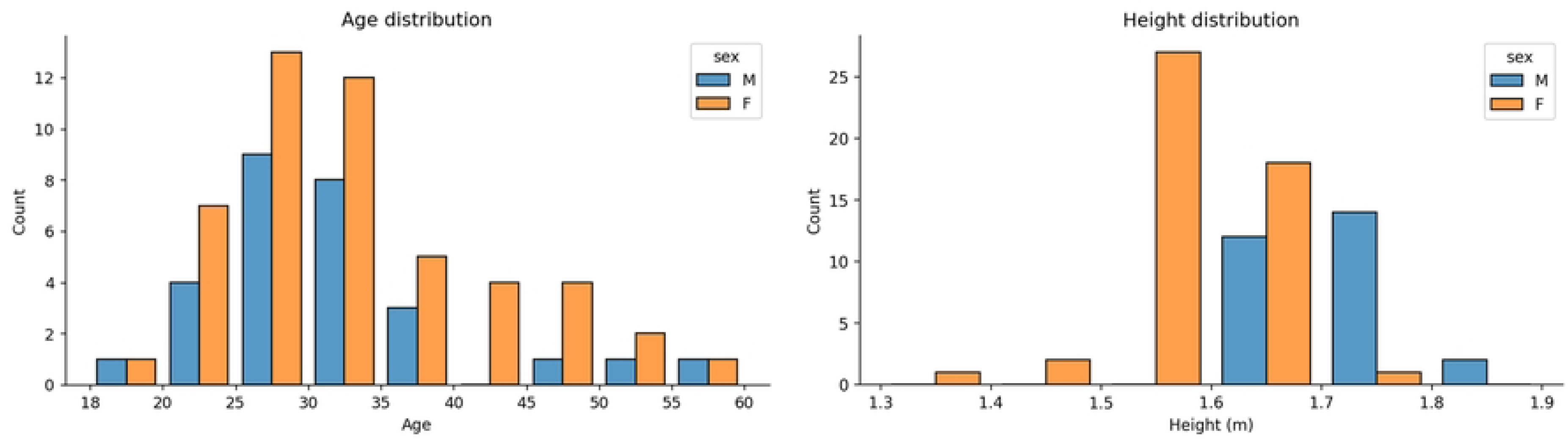
Age and height distribution of the included subjects, divided by sex.

Data were acquired on a 0.55 T Siemens Magnetom FreeMax using a 20-channel head coil. The entire acquisition protocol included *T*_2_-weighted, FLAIR and diffusion-weighted images in addition to the *T*_1_-weighted images used in this study. The *T*_1_-weighted images were acquired with an MPRAGE sequence with an echo time of 3.88 ms, a repetition time of 1700 ms, and a flip angle of 20 degrees. 192 sagittal slices were acquired, with a spatial resolution of 0.56 × 0.56 × 1 mm^3^. The image quality was evaluated using the MRIQC software [17], focusing on Signal-to-Noise Ratio (*SNR*) as defined by Dietrich et al. [18], as well as on the presence of ghosting and blurring artefacts, quantified through Shannon entropy of voxel intensities (entropy-focus criterion *efc*) [19] and on the white matter-to-maximum intensity ratio (*wm2max*)[20].

Images were preprocessed as follows: the original DICOM images were resampled to an isometric 1 mm^3^ spacing to match the voxel size of the MNI-152 template. Care was taken to maintain the images in the RPI orientation. Brains were then extracted using the skull-stripping algorithm HD-BET [21]. Background was cropped out in the three axial, sagittal and coronal orientations, removing the slices whose accumulated intensity was equal to zero and thus did not contain any brain information. Zero padding was then applied to the images, making their matrix size equal to 182 × 218 × 182, equivalent to the MNI-152 template size. The N4 bias field correction algorithm implemented in ANTs was applied [22]. Then, to maintain all the images with similar contrast, we matched each exam histogram to the average histogram of the entire group, using the “match histograms” function available in the scikit-image library in Python [23]. To generate this average histogram, we extracted histograms from the 76 selected images from our acquired data, after removing zero-valued background pixels and normalising all image intensities between 0 and 1. Each extracted histogram was binned into 255 bins, and we averaged them to obtain the reference histogram. Histogram matching was performed only on non-zero pixels, which were intensity-normalised between 0 and 1. We then restored these images to their original intensity range after histogram matching. Finally, the pixel intensities were adjusted so that the average gray matter value was 100. For that purpose, images were segmented into gray matter (GM), white matter (WM) and cerebrospinal fluid (CSF) using FSL v6.6.1 FAST algorithm [24].

### Template construction

The template was constructed using an iterative registration procedure, similar to [5,25]. The general workflow is presented in Fig 2. The procedure has two main stages: a preliminary *A0* template is built by rigid-body registration of the complete batch of preprocessed brain images to the standard MNI-152 template (NLin6Asym); then non-linear registrations are applied iteratively. The preprocessed images were first subjected to rigid-body registration to align each exam into a common space, using the MNI-152 as the initial reference. Registration was implemented using MATLAB v2024b (MathWorks, Natick, USA), using a gradient descent optimisation to iteratively minimise the difference between the moving image and the reference image, with 7 degrees of freedom allowing for global translation, rotation and scaling. Once all images were set in a similar reference frame, they were averaged to obtain an initial *A0* template.

**Fig 2.**
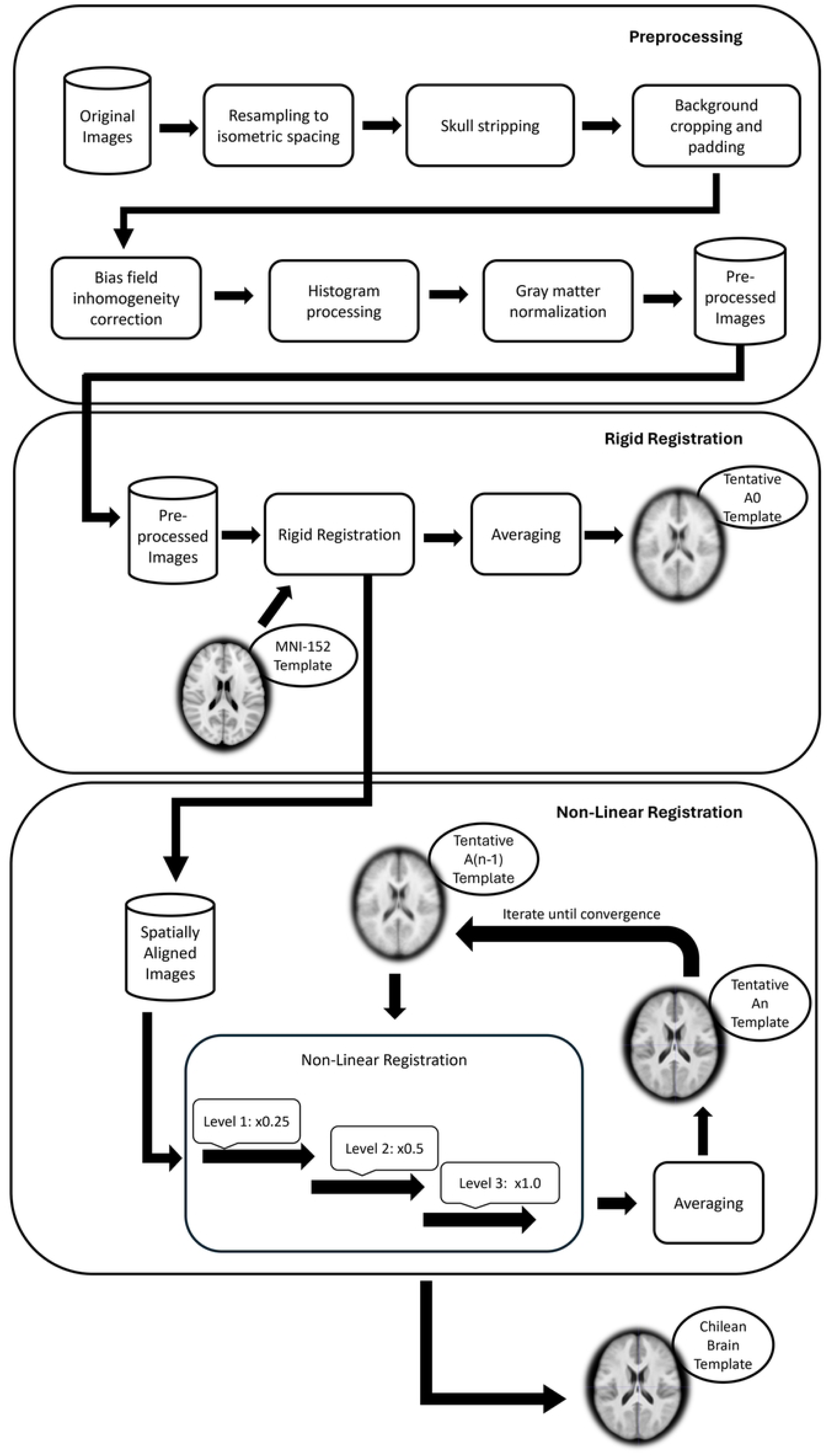
Pipeline for the template construction.

Iterative non-linear registration was then implemented to obtain a more precise template. At the *i*^th^ iteration, a template *Ai* is obtained by transformation of the previously spatially aligned images in the *A(i–1)* template. For the first iteration, the preliminary template *A0* was used. Non-rigid deformation was applied through symmetric normalisation (SyN) of the Advanced Normalization Tools (ANTs) [26,27]. The non-linear registration was implemented in three different resolution levels, starting with a coarse resolution registration at one-fourth of the image resolution, followed by a medium resolution registration at half the resolution, and a full resolution registration. Every registration level was performed for 50 cycles before moving to the next level. Once all exams were non-linearly registered, they were averaged to obtain a new *Ai* version of the template. As a result, this step produced a deformation field as a vector map describing each voxel’s displacement along the x, y, and z axes needed to meet the image registration criteria. The non-linear registration process was repeated until no differences were observed between consecutive iterations. We estimated the difference between iterations using the root mean square error (RMSE) function and continued iterating until the RMSE changed by less than 0.05 for three consecutive iterations. The resulting *An* image was presented as our final template, denominated CL76 in the rest of the work.

### Template evaluation

The obtained template was first compared with templates developed from different geographical areas: Canada (MNI152) [1,2], India (IN157) [5], China (CN200) [7], and Korea (KOR152) [28]. Anatomical description of the templates was obtained through: (1) the distance from the center of the anterior commissure (AC) to the center of the posterior commissure (PC) (i.e. the length of the AC-PC line); (2) the length of the whole brain, as the distance from the anterior pole to the posterior pole on the transverse plane across the AC-PC line; (3) the width of the whole brain determined by the distance of the line running through the middle point of the AC-PC line from the left pole to the right pole on the transverse plane; (4) the brain height measured as the distance from the superior pole to the inferior pole on the coronal plane. Along with these four features, we also considered the ratios width/length (W/L), height/length (H/L), and height/width (H/W).

The adequacy of the Chilean template for our local population was also evaluated by quantifying the deformation magnitude required, using the 23 subjects not included in the template construction as a testing group. The test group included 10 women (43.5%) and 13 men (56.5%), aged 18 to 60 years (average age 40.7 ± 13.7 years). The average height of test subjects was 1.62 ± 0.14 m for women and 1.74 ± 0.06 m for men, while the average weight of the women subjects was 71.77 ± 20.93 kg and 80.42 ± 10.87 kg for men. The study group had a mean body surface area of 1.89 ± 0.25 m^2^ (ranging from 1.44 to 2.52 m^2^).

We evaluated the deformation fields from the non-linear registration using the mean of the Jacobian determinant (*mJD*) and the mean deformation value (*mDV*) of the final template [7]. The *mJD*, as described in Eq (1), is obtained first by calculating the Jacobian determinant at each *i* voxel of the deformation field, expressed in the equation as |det(*J*^i^)|, where *J*^i^ is the Jacobian matrix in the *i* voxel of the deformation field and then calculating its average. Depending on its value, it indicates the type of deformation that occurred during registration: expansion when the Jacobian determinant is greater than 1, and shrinkage when it is less than 1.

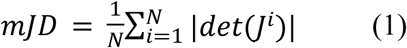

Eq (2) describes the *mDV*, which is obtained by adding the magnitude of deformation in the x, y, and z axes from the deformation map at each *i* voxel of the image and then averaging them.

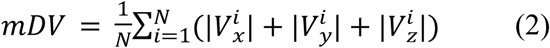

Statistical significance was evaluated with two-way ANOVA and F-test on the distribution of the deformation obtained over all subjects, considering α of 5%. Lilliefors tests were first run to assess the normality of deformation distributions across all subjects.

Beyond the global observation of deformation as such, to identify the main deformation loci in different cortical regions, we mapped the Automated Anatomical Labelling (AAL3) template onto each template included in this study [29]. The magnitudes of deformations *mDV* were averaged over each AAL region, and the ten areas with the most deformation were listed and contrasted qualitatively between templates.

The analyses were implemented on a server including an AMD EPYC 7443 processor setup, totalling 48 cores (96 threads) per processor. The system had 512 GB of RAM. The workload was distributed across 10 subroutines executing simultaneously, utilising 10 CPU cores and 50 GB of RAM. The resulting template is made available (https://github.com/SterenChabert/ChileanBrainTemplate).

## Results

Fig 3 shows example anatomical images of a subject, indicating that images obtained at 0.55T were of good quality. The image SNRs were satisfactory, with an average of 34.0 ± 9.8, close to the SNR observed in standard clinical settings using 1.5T. Artefact quantification remained low, with *efc* values of 0.52 ± 0.04, which is satisfactory. The impact of the low magnetic field strength was more pronounced in the white matter-to-maximum intensity ratio, with values somewhat smaller than usual (0.41 ± 0.03), whereas expected values are usually closer to 0.6 [20].

**Fig 3.**
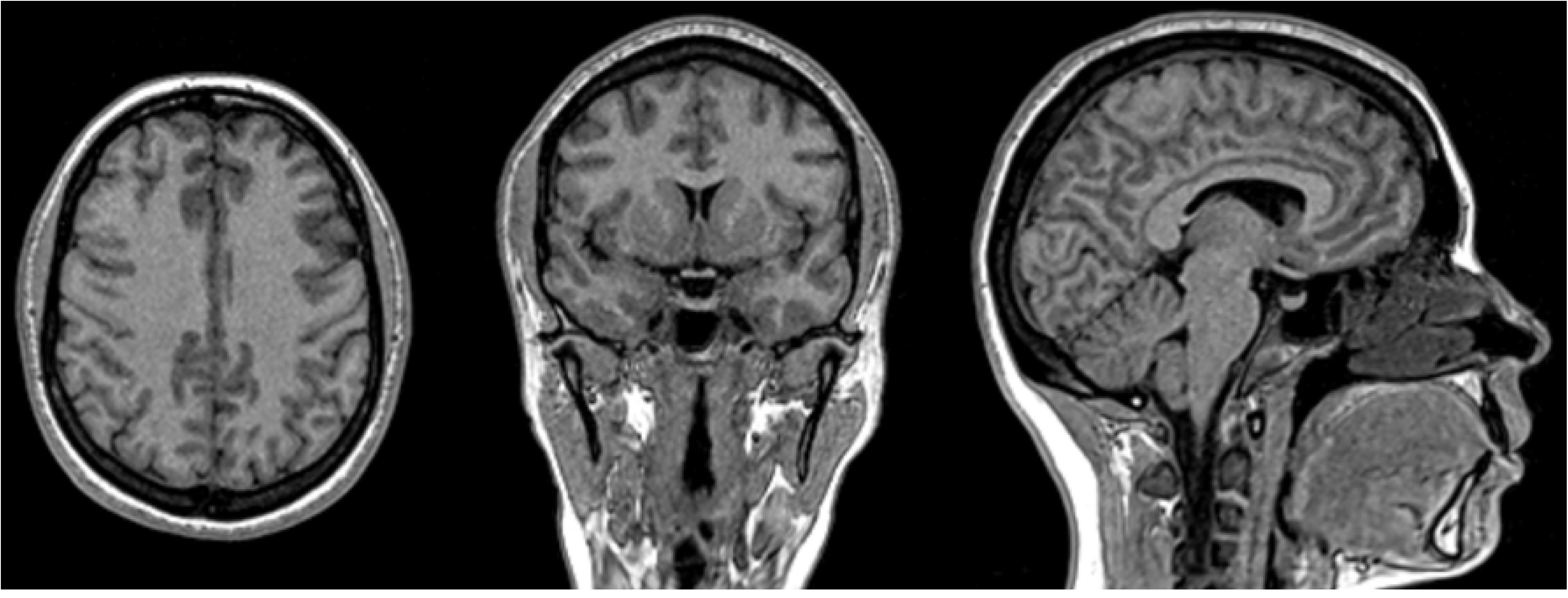
Example of T1-weighted images acquired at 0.55T. Axial (left), coronal (middle), and sagittal (right) T1-weighted slices obtained from a representative subject scanned at 0.55T. The images illustrate the overall structural detail achievable with the low-field system, showing adequate gray–white matter contrast, delineation of cortical sulci and deep nuclei, and preservation of midline structures. These acquisitions were part of the dataset used to build the Chilean brain template, highlighting the feasibility of using low-field MR technology for population-specific neuroimaging resources.

The evolution of the template built through iterations is presented in Fig 4, with the convergence process visible in Fig 5. The level of detail obtained in the final iteration is satisfactory, and its quality is visually equivalent to other published templates, as shown in Fig 6. The rigid registration procedure lasted a total of 36 minutes, and the non-rigid registration lasted an average of 133 minutes per subject in the *A*_0_ case, down to 16 minutes for the last template iteration.

**Fig 4.**
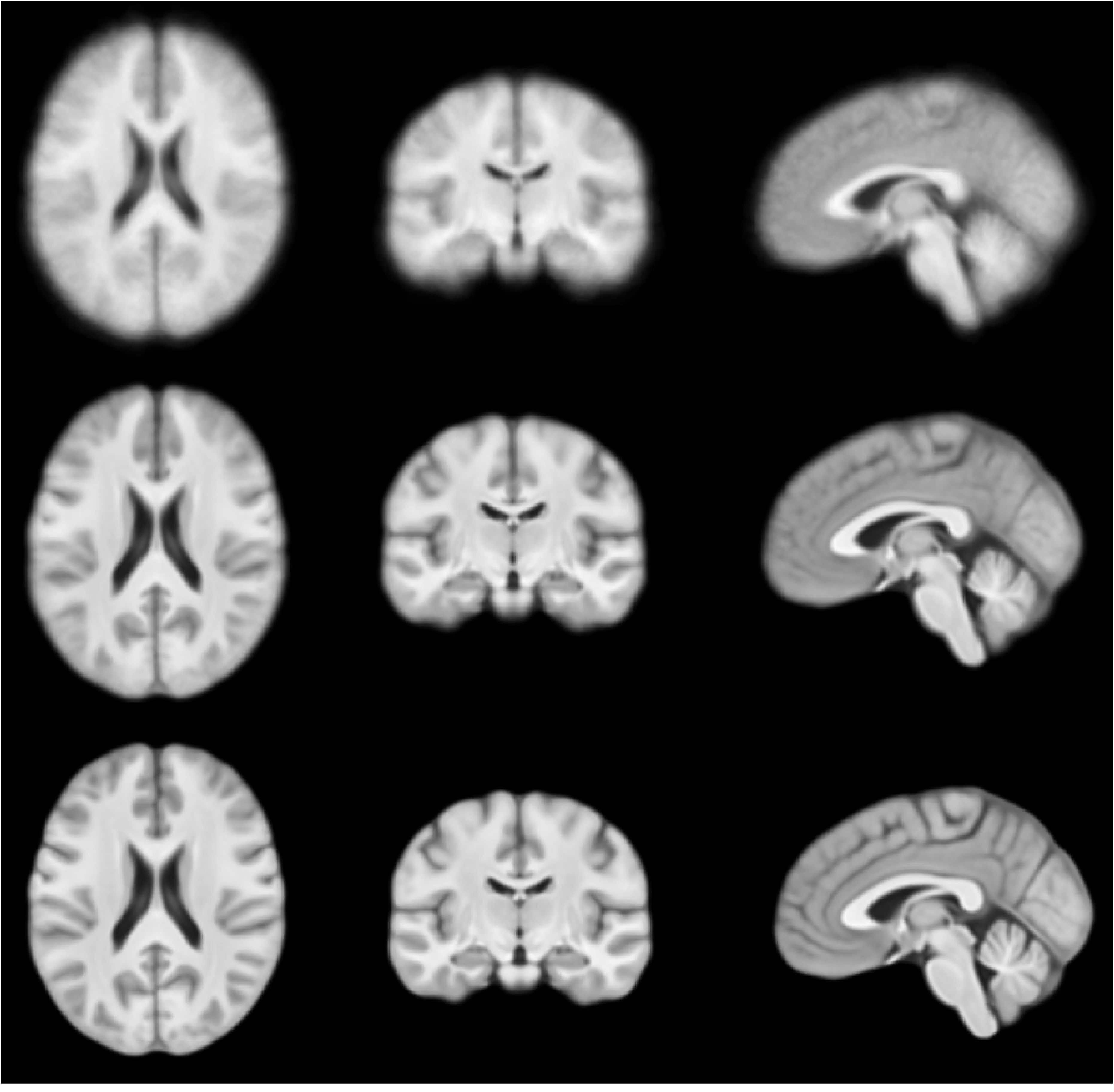
Evolution of the template across iterations. First row: axial, coronal and sagittal views of the template obtained in the initial round of iteration *A*_0_; central row: template obtained after the first iteration *A*_1_; bottom row: template obtained after the last iteration *A*_10_.

**Fig 5.**
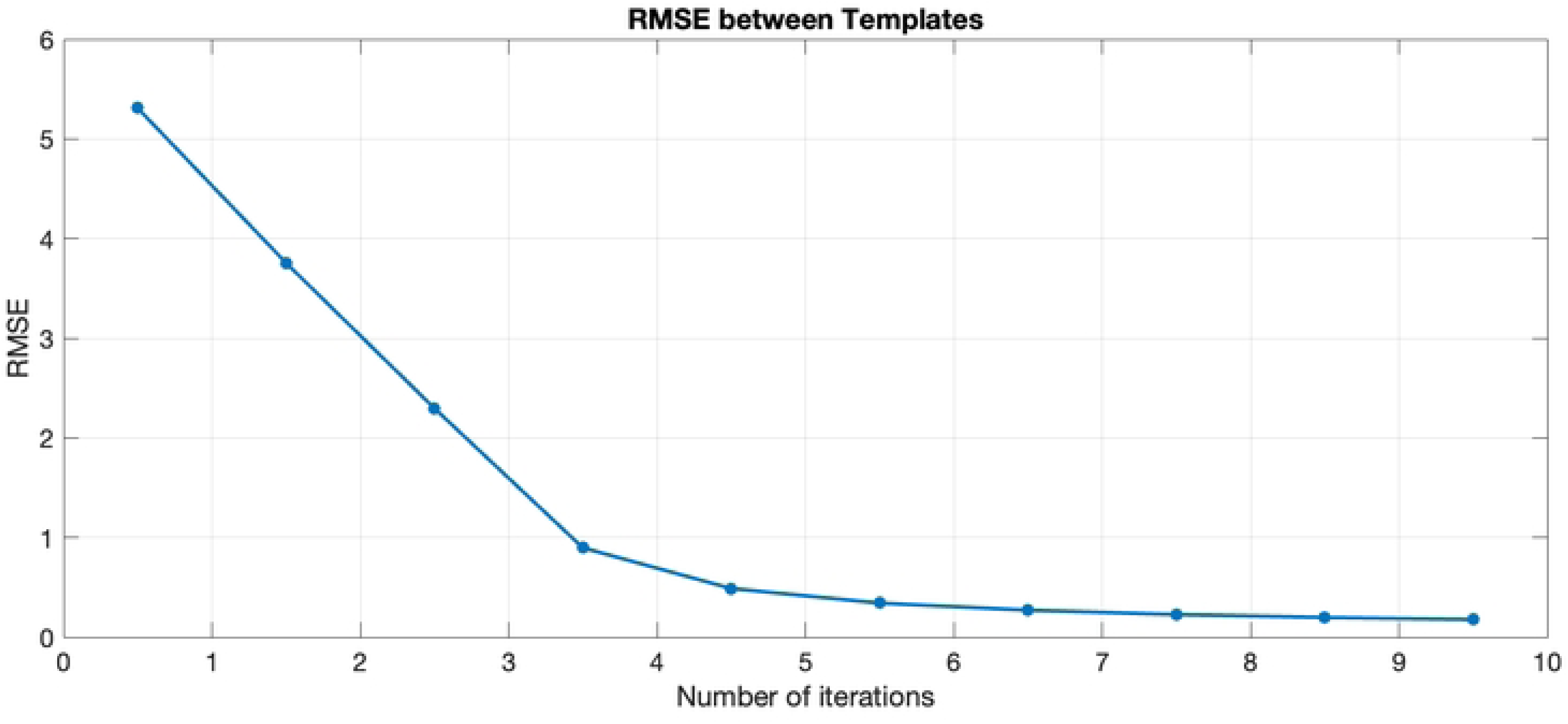
Convergence of template construction. Global root mean square error (*RMSE*) quantified between each iteration of template construction.

**Fig 6.**
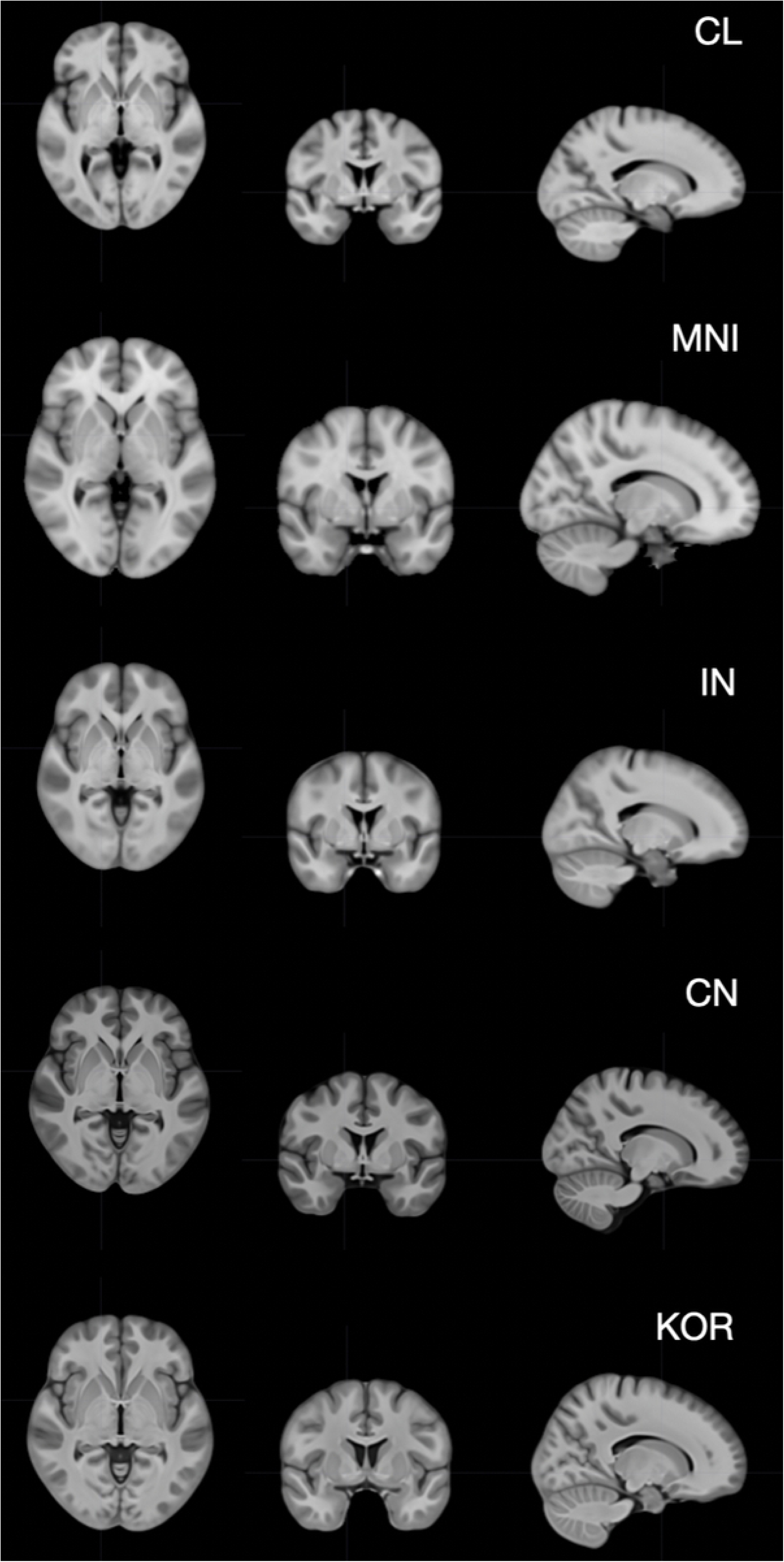
Templates compared in this work. Axial, coronal and sagittal views of the templates considered in this work, presented at a fixed scale, to visualize the differences in size and aspect ratio. From top row to bottom row: Chilean (CL), Canadian (MNI), Indian (IN), Chinese (CN) and Korean (KOR) templates.

Fig 6 shows the templates obtained from images in Canada, India, China, and Korea, as well as from our local data in Chile. Table 1 provides the quantitative description. As expected, templates from countries where the population tends to be higher, such as Canada, present a bigger template size: 179 × 137 × 118 mm^3^. In contrast, the Chilean template measures 13 × 5 × 18 mm^3^, smaller than the MNI one, corresponding to a relative difference of 4% to 15%. In terms of global size, the templates closest to the Chilean one are those from Korea or India, but their three-dimensional proportions are not similar. The height of the Chilean template is the smallest one: only 10 cm, while the template height from Canada is close to 12 cm. Beyond the sheer differences in length, width and height, the hull proportions for each template vary: for instance, the Chinese and Korean templates are “rounder”, with a length/width ratio close to 1.18, while the MNI template from Canada is “lengthier” in the antero-posterior direction with a length/width ratio of 1.31. The Chilean template has the largest width/height and length/height ratios (1.31 and 1.65, respectively) relative to the smallest height measured among all templates.

**Table 1.** Size comparison between the Chilean template and other templates.

|  | **CL76** | **MNI152** | **IN157** | **CN200** | **KOR152** |
|---|---|---|---|---|---|
| AC-PC (mm) | 27.1 | 27.8 | 26.0 | 26.0 | 24.9 |
| Length (mm) | 165.8 | 179.0 | 166.3 | 164.5 | 160.4 |
| Width (mm) | 132.0 | 137.0 | 129.5 | 140.0 | 134.7 |
| Height (mm) | 100.4 | 118.0 | 115.1 | 114.7 | 113.2 |
| Length/width ratio | 1.26 | 1.31 | 1.28 | 1.18 | 1.19 |
| Width/height ratio | 1.31 | 1.16 | 1.13 | 1.22 | 1.19 |
| Length/height ratio | 1.65 | 1.52 | 1.44 | 1.43 | 1.42 |

The improved adequacy of the Chilean template to Chilean subjects is quantified through the more minor net deformation needed (*mJD*), in contrast to the one needed using the MNI Canadian template, shown in Fig 7. The average net deformation is significantly smaller when using the Chilean template (p = 0.010), whereas the effect of the MNI template is more pronounced than that of most subjects visible on the right side of Fig 7. There is no significant difference in average deformation obtained with the Chilean template or any one of the Asian templates, Indian, Chinese or Korean (p = 0.77, 0.43 and 0.84, respectively). The variance of the required deformation was smaller with the Chilean template than with the Chinese and Korean templates (p = 0.015 and 0.027, respectively), indicating that a local proposal is globally more adequate as a structural reference. All deformation distributions were normally distributed. In all cases but the Canadian template, the average magnitude of deformation (*mDV*) did not vary significantly with the height of the subject, as shown in Fig 8.

**Fig 7.**
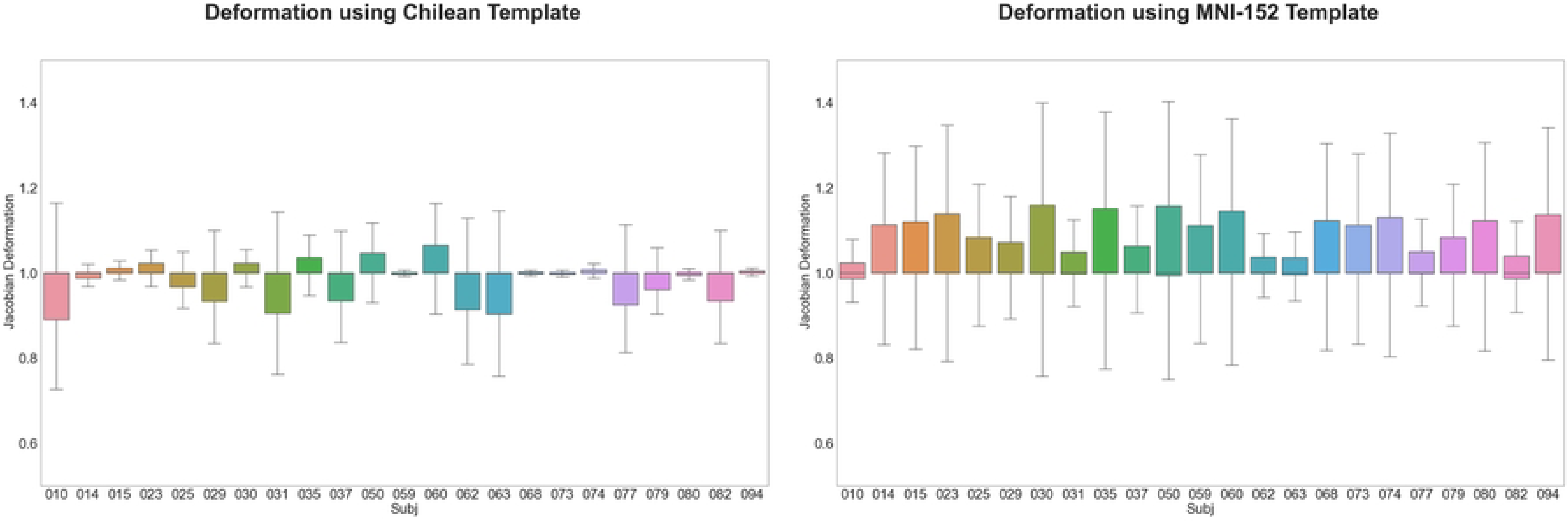
Deformation needed to register the test subjects onto the Chilean and Canadian templates. Comparison of the magnitude of deformation needed for the 23 subjects used for testing, onto the Chilean template (left) and the Canadian template (right). Quantification is expressed using the mean of the determinant of the Jacobian matrix; the *x* axis presents each one of the different subjects.

**Fig 8.**
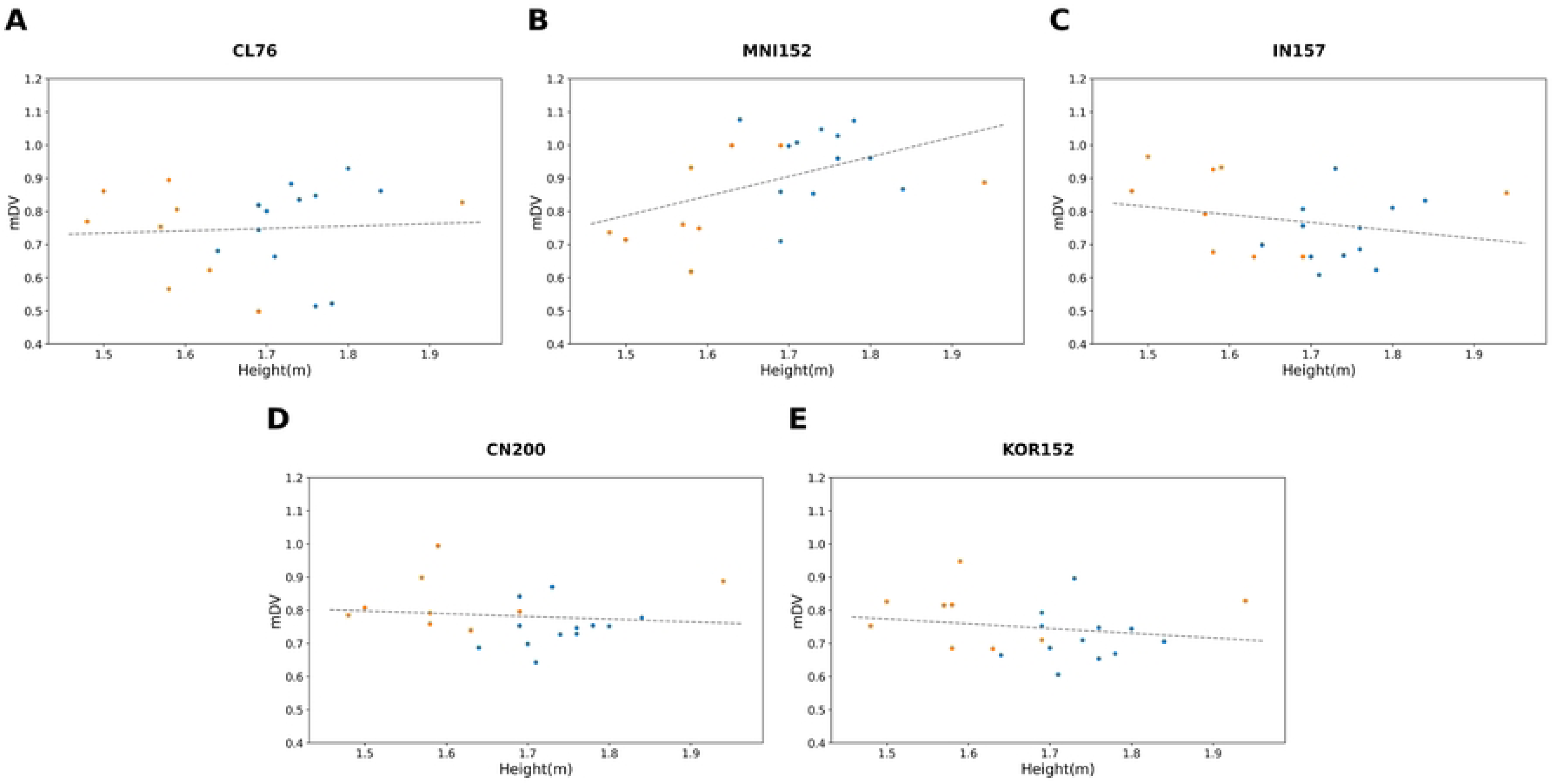
Deformation (mDV) in relation to the height of the Chilean test subjects for each template. (A) Chilean template (R^2^ = 0.004, Pearson correlation coefficient of 0.06 with p = 0.80). (B) Canadian template (R^2^ = 0.24, Pearson correlation coefficient of 0.49 with p = 0.03). (C) Indian template (R^2^ = 0.06, Pearson correlation coefficient of –0.24 with p = 0.30). (D) Chinese template (R^2^ = 0.01, Pearson correlation coefficient of –0.12 with p = 0.62). (E) Korean template (R^2^ = 0.04, Pearson correlation coefficient of –0.19 with p = 0.41). Blue dots indicate male subjects and orange dots indicate female subjects.

Beyond the global dimension or magnitude of deformation, Table 2 provides an approximation of which ten AAL areas are most deformed relative to each template. The point is not to interpret a specific area and/or function that would be more represented in one case or the other, but to emphasise that the locus of non-linear deformation associated with each template differs within the brain. Of the 10 AAL regions associated with the most deformation magnitudes in the Chilean case, none are in common with the 10 most deformed AAL areas in the Canadian case, while seven are also found in the Korean template (Posterior orbital gyrus (R), IFG pars orbitalis (L), Lobule VI of vermis, Posterior orbital gyrus (L), Heschl’s gyrus (R), Temporal pole: superior temporal gyrus (L) and Lateral orbital gyrus (R)), or even 8 in case of considering as well the contra-lateral area Rolandic operculum (L). In a similar manner, 6 out of 10 areas are also found using the Chinese template (Posterior orbital gyrus (R), IFG pars orbitalis (L), Posterior orbital gyrus (L), Heschl’s gyrus (R), Temporal pole: superior temporal gyrus (L) and Lateral orbital gyrus (R)), or even 8 in case of considering as well the contralateral area Rolandic operculum (L) and Heschl’s gyrus (L). In the Indian template, only 4 out of the 10 regions are found associated with the most deformation in the Chilean case. In a complementary way, 7 of the 10 areas listed as most deformed in the Canadian template are found only in the MNI case, and not in the other proposed templates, emphasising the underlying structural differences between templates, and the more pronounced difference in the Canadian case with respect to the Asian and Chilean templates.

**Table 2.** Most deformed regions. Ten AAL3 regions with the highest mean deformation value (mDV) when registering the test subjects onto each template. L, left; R, right; IFG, inferior frontal gyrus.

| **CL76** |  | **MNI152** |  |
|---|---|---|---|
| **AAL area** | **mDV** | **AAL area** | **mDV** |
| Posterior orbital gyrus (R) | 3.885 | Substantia nigra, pars reticulata (L) | 4.050 |
| IFG pars orbitalis (L) | 3.628 | Medial orbital gyrus (L) | 3.990 |
| IFG pars orbitalis (R) | 3.611 | Precentral gyrus (R) | 3.943 |
| Lobule VI of vermis | 3.599 | Substantia nigra, pars reticulata (R) | 3.922 |
| Rolandic operculum (R) | 3.495 | Lobule I, II of vermis | 3.897 |
| Posterior orbital gyrus (L) | 3.482 | Lateral orbital gyrus (L) | 3.774 |
| Heschl's gyrus (R) | 3.473 | Posterior orbital gyrus (L) | 3.745 |
| Temporal pole: superior temporal gyrus (L) | 3.459 | Medial orbital gyrus (R) | 3.691 |
| Lateral orbital gyrus (R) | 3.428 | Heschl's gyrus (L) | 3.679 |
| Temporal pole: superior temporal gyrus (R) | 3.391 | Precentral gyrus (L) | 3.638 |
| **IN157** |  | **CN200** |  |
| **AAL area** | **mDV** | **AAL area** | **mDV** |
| Heschl's gyrus (R) | 3.926 | Posterior orbital gyrus (R) | 3.872 |
| Lobule VIII of vermis | 3.771 | Posterior orbital gyrus (L) | 3.831 |
| Lobule VIIB of cerebellar hemisphere (R) | 3.658 | Temporal pole: superior temporal gyrus (L) | 3.757 |
| Crus II of cerebellar hemisphere (R) | 3.633 | Heschl's gyrus (R) | 3.749 |
| Lobule VII of vermis | 3.631 | Heschl's gyrus (L) | 3.733 |
| Lobule VI of vermis | 3.602 | Rolandic operculum (L) | 3.719 |
| IFG pars orbitalis (L) | 3.591 | Lobule X of vermis | 3.613 |
| Posterior orbital gyrus (R) | 3.551 | IFG pars orbitalis (L) | 3.604 |
| Lobule X of vermis | 3.545 | Lateral orbital gyrus (R) | 3.565 |
| Lobule IX of vermis | 3.510 | Temporal pole: middle temporal gyrus (L) | 3.524 |
| **KOR152** |  |  |  |
| **AAL area** | **mDV** |  |  |
| Posterior orbital gyrus (R) | 3.651 |  |  |
| Heschl's gyrus (R) | 3.619 |  |  |
| IFG pars orbitalis (L) | 3.531 |  |  |
| Posterior orbital gyrus (L) | 3.468 |  |  |
| Lateral orbital gyrus (R) | 3.437 |  |  |
| Temporal pole: superior temporal gyrus (L) | 3.424 |  |  |
| Crus II of cerebellar hemisphere (L) | 3.418 |  |  |
| Rolandic operculum (L) | 3.384 |  |  |
| Lobule VI of vermis | 3.365 |  |  |
| Crus II of cerebellar hemisphere (R) | 3.356 |  |  |

## Discussion

To our knowledge, this is a one-of-a-kind template not only for the population it represents but also due to the low magnetic field strength used to obtain the images, 0.55 T. A slight effect of image quality differences, such as contrast or spatial resolution, associated with a low *B*_0_ could have affected our result, but, in our estimation, this possible variation associated to *B*_0_ should be smaller than the main effect of broader template variation. This possible effect calls for further work to determine it more clearly.

The different templates published in the literature [2,5,7,28] include subjects using a variety of criteria, most commonly healthiness defined in the absence of any diagnostic or pathological condition, or defined according to performance on cognitive evaluations. We decided to take a mixed approach: not only the absence of a diagnosis but also review by expert neuroradiologists, which would guarantee normal anatomy. We were surprised by the high incidence of anatomical findings that excluded up to 25% of our data—most of which were not relevant to pathological definitions but were visible enough to warrant excluding these images from constructing a “normal” reference template. As a consequence, one limitation of this work is the limited sample size used for template construction. Yang et al. [7] recommended a sample size of 200 for brain template elaboration, and emphasised that a small sample size would lead to higher brain template variability. The comparison was made here with templates elaborated with a size double that of ours: 152 for the Canadian template, 152 for the Korean one, 157 for the Indian one, and 200 for the Chinese template. Yet the template whose structure appeared most similar to the Chilean one is the Chinese one, suggesting that the differences observed between templates go beyond differences in sample size used in their elaboration. It is worth noting as well that the median value for template construction found in the literature is 43 [10].

A limitation of this study is that the test group was small and composed of subjects excluded for radiological findings. Their anatomy therefore departs from that of the healthy population the template represents. Nevertheless, this situation mirrors the clinical use of templates, which are routinely applied to patients with anatomical variants or incidental findings. Future work should extend the evaluation to a larger cohort, including healthy subjects not used for template construction, and assess the impact on downstream analyses.

In this one template, adults from 18 to 60 years old are represented, considered as being neither “too young” nor “too old”, beyond the active discussion associated with normal “regular” ageing and possible burst of ageing at different moment [30]. Volunteers were recruited only in the capital city, which does not fully represent the diversity of the population of an American country 4000 km long. From a genetic perspective, the Chilean population is primarily the result of admixture between European settlers and the Indigenous peoples of Chile, especially the Mapuche and Aymara, with a minor but widespread African component [31,32]. Genetic studies show a strong asymmetry: most mitochondrial DNA (inherited maternally) is of Indigenous origin (up to 84% in Santiago), while the Y-chromosome (inherited paternally) is predominantly European (about 70%). Any population heterogeneity is hard to represent accurately, but the challenge is greater when population origins are broader.

Interestingly, the Asian templates appear closer to the Chilean one than to the Canadian template, which is based on the North American continent. This observation is coherent with the known average height of the population: in Chile the average height of men was 173.1 cm in 1980 [33] — even though these are not the most recent data, 1980 corresponds to the closest common year of publication of the international data of interest here. These values are closer to the 170.2 cm measured for Korean men [34] than to the 179.6 cm measured in Canada [33]. The Chilean template’s proximity to Asian templates may reflect that South American populations retain a larger relative proportion of Native American genetic ancestry than those in North America. In a recent survey, 67% of the Canadian population declared themselves as “white”, 27% as “minority” (South Asian, Chinese or other) and 6.0% as Indigenous [35]. The founding Indigenous ancestry originates from ancient East Asian populations that entered via Beringia, leading to subsequent population splits and migrations that have shaped the current genetic landscape [36]. Yet this is only a broad description for possible interpretation, and this work does not pretend to replace more detailed genetic studies, especially in a continent, America, intrinsically constituted by immigration from different places in the world.

The need for accurate, population-specific brain templates is well established. Using templates that do not match the study population introduces measurable distortions in morphometric estimates and registration accuracy in magnetic resonance imaging [5,7,10–13,37]. Such distortions are not merely technical but can have direct clinical implications: studies have shown that template-induced biases may alter lesion localisation, compromise surgical planning, and affect group-level inferences, ultimately influencing patient care [37,38]. Importantly, population-matched templates have been demonstrated to yield higher statistical power and stronger effect sizes when detecting disease-related changes, such as gray matter atrophy in Alzheimer’s disease [12,39]. However, constructing appropriate templates remains challenging. Building representative atlases requires access to large, well-characterised cohorts, and even then, a single template may not account for relevant comorbidities or the heterogeneity inherent in diverse populations. Future work should address the granularity at which sub-populations should be defined and determine at what point anatomical generalisations can be considered valid without compromising accuracy.

## Conclusion

This study shows that recent 0.55T MR scanners can provide images of sufficient quality for effective template building. To the best of our knowledge, this is the first brain template generated in South America; however, we do not claim to represent the entire continent—only a specific population from Chile. This work highlights the need for broader representation across diverse populations, particularly to counterbalance the current over-representation of individuals from high-income countries with established research and imaging infrastructures. As with mannequins, the concept of a “one-size-fits-all” brain template has inherent limitations and may not optimally capture the anatomical variability relevant to different populations or study objectives.

Our findings reinforce the need to develop population-specific clinical templates. Reliance on non-representative atlases, such as MNI152, may introduce systematic biases in diagnosis, lesion localisation, and surgical planning. Locally derived templates can provide more accurate and clinically relevant neuroimaging evaluations, ultimately improving patient care in diverse populations. For Chile, Asian templates would likely offer a more suitable alternative than the widely used MNI152 from Canada. Still, there remains a need to better define and quantify the impact of using one template versus another, especially in terms of downstream analyses and interpretation.

## Data Availability

The resulting template is made available (https://github.com/SterenChabert/ChileanBrainTemplate).

## Supporting information

**S1 Table. Radiological findings associated with the exclusion of volunteers.** Findings reported by neuroradiologists for each of the 23 excluded volunteers.

